# Trust in physicians as a mediator of the relationship between person-centered care and medication adherence in patients undergoing hemodialysis: A cross-sectional study

**DOI:** 10.1101/2025.01.02.25319883

**Authors:** Yusuke Kanakubo, Ryohei Inanaga, Tatsunori Toida, Tetsuro Aita, Mamiko Ukai, Atsuro Kawaji, Takumi Toishi, Masatoshi Matsunami, Yu Munakata, Tomo Suzuki, Tadao Okada, Noriaki Kurita

**Affiliations:** Tessyoukai Kameda Family Clinic Tateyama, Tateyama-City, Chiba, Japan; Division of Clinical Epidemiology, Research Center for Medical Sciences, The Jikei University School of Medicine, Minato-ku, Tokyo, Japan; Department of Nephrology, Shin-Yurigaoka General Hospital, Kawasaki-City, Kanagawa, Japan; Department of Clinical Epidemiology, Graduate School of Medicine, Fukushima Medical University, Fukushima-city, Fukushima, Japan; School of Pharmaceutical Sciences, Kyushu University of Medical Science, Nobeoka-city, Miyazaki, Japan; Department of General Internal Medicine and Family Medicine, Fukushima Medical University, Fukushima City, Fukushima, Japan; Department of Nephrology, Kameda Medical Center, Kamogawa-City, Chiba, Japan; Chikuseikai Munakata Clinic, Shinjuku-ku, Tokyo, Japan; Munakata Clinic, Mobara-city, Chiba, Japan; Department of Innovative Research and Education for Clinicians and Trainees (DiRECT), Fukushima Medical University Hospital, Fukushima-city, Fukushima, Japan

## Abstract

**Rationale & Objective:** Person-centered care (PCC) and trust in physicians influence medication adherence (MA) among patients undergoing dialysis. However, the mechanisms linking PCC to MA, particularly the mediating effect of trust in physicians, remain unclear. This study aimed to investigate the interrelationships between PCC, trust in physicians, and MA.

**Study Design:** Multicenter cross-sectional study.

**Setting & Participants:** Japanese adults receiving outpatient hemodialysis at six dialysis centers.

**Exposures:** PCC was assessed using the 13-item Japanese Primary Care Assessment Tool-Short Form, which included longitudinality and coordination of care. Trust in physicians was measured using the five-item Wake Forest Physician Trust Scale.

**Outcome:** MA was measured using the 12-item Adherence Starts Knowledge (ASK-12) scale.

**Analytical Approach:** General linear models were used to examine the relationships between PCC, trust in physicians, and MA. Mediation analysis determined the extent to which trust in physicians mediated the PCC-MA relationship.

**Results:** In total, 483 patients were analyzed. High-quality PCC was associated with lower barriers to MA in a dose-response manner across JPCAT-SF quartiles compared to no usual source of care (Q1: −3.90 (95% CI: −5.95 to −1.75); Q2: −4.78 (95% confidence interval [CI]: −7.02 to −2.69); Q3: −4.69 (95% CI: −6.96 to −2.58); Q4: −4.95 (95% CI: −7.21 to −2.62). Trust in physicians partially mediated this relationship in a dose-response pattern, with the proportion of the indirect effect increasing from 16.1% (95% CI: 4.5 to 33.8%) in Q2 to 33.3% (95% CI: 17.4 to 65.5%) in Q4. Similar findings were observed for PCC subdomains.

**Limitation:** Possible reverse causation.

**Conclusions:** High-quality PCC was associated with MA, with trust in physicians playing a key mediating role. Strategies to enhance MA in patients undergoing hemodialysis should incorporate multidimensional PCC approaches, focusing on building trust and strengthening continuity and coordination of care.

## Introduction

Poor medication adherence is frequently reported among patients undergoing hemodialysis, who often require 10 to 20 tablets daily to manage complications related to chronic kidney disease (CKD) and comorbidities. [1] Studies have consistently reported non-adherence rates >50%, with some estimates as high as 98.6%. [2–6] The impact of non-adherence in patients on dialysis is underscored by inadequate management of CKD-related complications and an associated increase in mortality rates. [2,7] A multidimensional approach to improving medication adherence has been proposed, incorporating not only medication-related factors (simplifying regimens) [6,8] and patient-related factors (enhancing health literacy) [9,10] but also strengthening physician-patient communication. [8] To achieve this, establishing effective physician-patient communication through person-centered care (PCC) has been advocated for patients with CKD. [8,11,12] However, despite its theoretical significance, empirical evidence supporting PCC remains limited.

PCC integrates not only evidence-based, disease- and organ-specific approaches but also an understanding of what matters most to individual patients, facilitating shared decision-making in their care. [12,13] When addressing treatment non-adherence, the PCC approach emphasizes understanding individual patient preferences, goals, and values, particularly for nephrologists. [12] Qualitative studies have suggested that key PCC elements, such as care continuity, [10] needs-driven practice rather than strict adherence to practice guidelines, [11] and coordination (ensuring consistency in treatment recommendations), [14,15] may contribute to improved medication adherence. However, these findings have been primarily limited to hypothesis generation.

In addition, limited studies have explored how PCC affects doctor-patient interactions in patients with kidney disease and, in turn, affects medication adherence. Theoretically, this interaction acts as a critical mediator between PCC and self-care behaviors such as medication adherence, [16] with trust in physicians playing a central role. [17] A study of patients with non-dialysis-dependent CKD demonstrated the association between PCC and trust in nephrologists. [18] In contrast, patients undergoing hemodialysis often perceive their medical facility as a usual source of care (USC), providing PCC for routine health issues as they visit the facilities three times a week. [19] Our previous findings indicated that trust in physicians facilitates medication adherence among patients on hemodialysis. [9] Examining specific PCC aspects enhancing medication adherence through trust in physicians may help identify tailored care strategies to optimize PCC, both within and beyond the physician-patient relationship.

This study aimed to address these gaps by focusing on Japanese patients undergoing hemodialysis. The specific objectives were: (1) to explore the interrelationships among PCC, trust in physicians, and medication adherence and (2) to examine the mediating role of trust in physicians in the relationship between PCC and medication adherence using mediation analysis.

## Methods

### Patient and Public Engagement Summary

Patients and/or the public were not involved in this study.

### Study Design and Participants

This multicenter, cross-sectional study was conducted at six medical facilities providing outpatient hemodialysis services. The study adhered to the principles of the Declaration of Helsinki and received ethical approval from the Ethical Review Board of Fukushima Medical University (Approval number: ippan2021-292). The inclusion criteria were: (1) adult patients with kidney failure undergoing hemodialysis or a combination of hemodialysis and peritoneal dialysis, (2) patients who regularly attended the participating facility for dialysis treatment, and (3) those who were capable of completing a questionnaire. Patients were excluded if they (1) did not have regular medication prescriptions or (2) suffered from severe dementia or complete blindness. All participants provided written informed consent and completed paper-based questionnaires. Data collection was conducted between April 2022 and February 2023.

## Measures

### Medication Adherence

The primary outcome measured was medication adherence, assessed using the Japanese version of the Adherence Starts with Knowledge (ASK)-12 scale. [20,21] This 12-item questionnaire evaluates medication adherence across three domains: inconvenience/forgetfulness, treatment beliefs, and behavior (Supplementary Item 1).

Each item is rated on a five-point scale, with higher scores indicating greater adherence difficulties. Items 4LJ7 were reverse-scored to maintain consistency with the other items. The total ASK-12 score, ranging from 12 to 60, was calculated by summing all item scores. The ASK-12 scale has demonstrated validity and internal consistency (coefficient alpha: 0.75) [20]. The criterion validity of the Japanese version was previously established in studies involving patients with asthma. [21,22]

### Patient Experience of Person-Centered Care

The primary exposure variable was the perceived quality of (PCC) by patients, measured using the 13-item Japanese Primary Care Assessment Tool-Short Form (JPCAT-SF). [19,23] The full version of the JPCAT-SF is an adaptation of the Primary Care Assessment Tool to Japanese practice and evaluates PCC, an essential element of primary care. [24,25] The instrument serves as a healthcare quality indicator by capturing the experiences of patients during the PCC process. [25,26] The JPCAT-SF includes the following six domains similar to the JPCAT: first contact (2 items), longitudinal (2 items), coordination (3 items), comprehensiveness (2 items each for services available and services provided), and community orientation (2 items). The JPCAT-SF has demonstrated good internal consistency reliability (coefficient alpha >0.76 for total and each domain scores) and strong criterion validity. [23] Details of the JPCAT-SF items and domains are provided in Supplementary Items 2 to 3.

Patients answered “yes” or “no” to the initial JPCAT-SF question: “Is there a doctor that you usually go to if you are sick or need advice about your health?” Patients responding “yes” were categorized as having a USC. Subsequently, JPCAT-SF scores were calculated for patients with USCs. Each item was rated on a five-point Likert scale, and domain scores were computed on a scale from 0 to 100 based on the established algorithm. [23] The overall JPCAT-SF score was the average of all domain scores, with higher scores indicating higher PCC quality.

### Trust in Physicians

Trust in physicians was analyzed as a mediator using the five-item Japanese version of the Wake Forest Trust in Doctors Generally scale. [9,27,28] This scale consists of five items rated on a five-point Likert scale, where patients indicate their agreement with each statement, ranging from “strongly disagree” (1 point) to “strongly agree” (5 points). Scores for negatively worded items were reversed, and the total score was scaled from 0 to 100. The construct validity and internal consistency of the scale were verified (coefficient alpha: 0.88). [27]

### Covariate Measurements

Confounding variables were selected based on existing literature and expert medical knowledge, focusing on factors likely to influence PCC, trust in physicians, and medication adherence. These variables included age, sex, dialysis duration, smoking history, level of education, household income, comorbid conditions (such as hypertension, diabetes, cardiovascular disease, liver disease, depression, and dementia), the total number of antihypertensive medication classes, the number of prescribed phosphate binders, and health literacy as measured by the Functional, Communicative, and Critical Health Literacy scale. [29] Detailed definitions of each covariate are provided in Supplemental Item 4. The questionnaire was distributed at each facility, and participants were instructed to complete it.

### Statistical Analysis

All statistical analyses were conducted using Stata/SE version 18 (StataCorp, College Station, TX). Patient characteristics were summarized using median and interquartile range (IQR) for continuous variables and frequencies with percentages for categorical variables. General linear models were employed to analyze the relationship between trust in physicians and PCC (Supplementary Figure 1, Analysis 1), as well as the association between ASK-12 scores and PCC, with and without considering trust in physicians (Supplementary Figure 1, Analysis 2). The JPCAT total score was classified into five multi-categories. [19] The response of no USC was set as the reference category. The remaining four categories were determined according to the JPCAT total score, with the number of people in each category as equally distributed as possible.

Mediation analyses were conducted to determine whether trust in physicians mediated the relationship between PCC and medication adherence. These analyses estimated the independent variable effect (PCC, the direct effect) after removing the mediator effect (trust in physicians), and the effect explained by the mediator (indirect effect).

Assuming causality, the indirect effect represents PCC influence on medication adherence mediated through trust in physicians, whereas the direct effects are those not mediated. The mediation analyses are outlined in Figure 1. Indirect effects were formally inferred using percentile bootstrap confidence intervals to assess whether trust in physicians mediates the relationship between PCC quality and medication adherence. [30] Indirect effects are deemed statistically different from zero if the confidence interval does not span zero.

**Figure 1.**
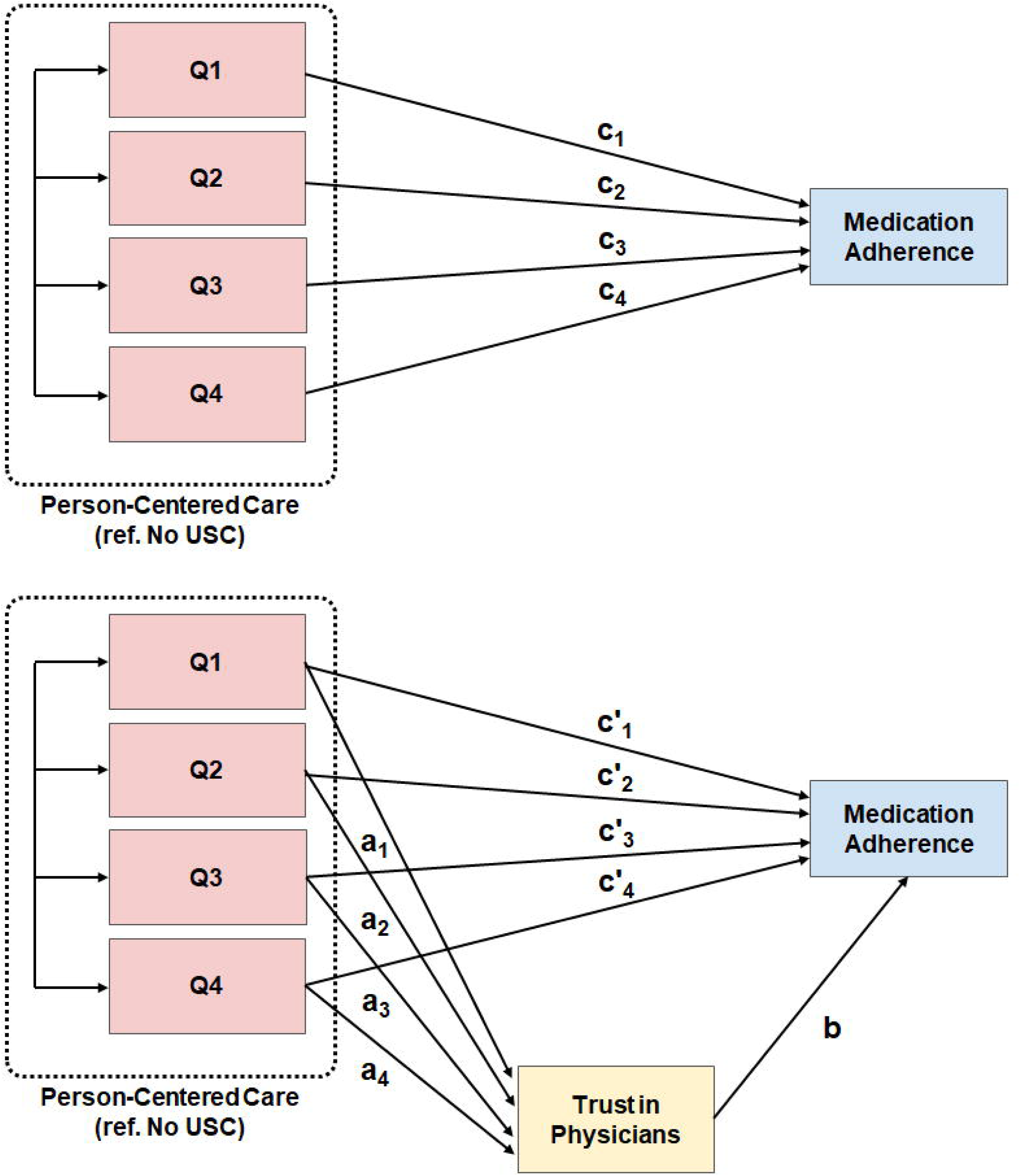
Mediation analysis. The mediation analysis was conducted in three steps: First, medication adherence was regressed on PCC and its covariates, excluding trust in physicians. The resulting coefficients for PCC in this step, denoted as cLJ to cLJ, represent the total PCC effect on medication adherence (the effect before accounting for mediation by trust in physicians, Upper half of the figure). Second, trust in physicians, the mediator was regressed on PCC and covariates. PCC coefficients in this regression are denoted as aLJ to aLJ. Third, medication adherence was regressed on PCC, trust in physicians, and covariates. The coefficient for trust in physicians in this model is b, while PCC coefficients are denoted as c’LJ to c’LJ, which represent the direct effect of PCC on medication adherence. The indirect effect of PCC on medication adherence, mediated through trust in physicians, is calculated as the product aLJ × b to aLJ × b. The total effect (cLJ) is the sum of the direct effect (c’LJ) and the indirect effect (aLJ × b), where i = 1 to 4. (Bottom half of the figure)

Similar to the main analysis, the mediating role of trust in physicians in the relationship between the JPCAT-SF subdomain score (classified into five categories with no USC as a reference) and medication adherence was separately analyzed using general linear models adjusted for the same covariates. Twenty imputations were conducted using multiple imputations with chained equations, assuming the data were missing at random. [31] The aforementioned bootstrap samples were first generated from the raw data, followed by multiple imputations of the missing values in each of the bootstrap samples. [32] A total of 1,000 re-samplings were conducted. Confidence intervals (CIs) for model parameters were estimated using percentile-based bootstrapping. This approach enabled the calculation of valid CIs, even in the presence of heteroscedasticity in the model residuals. [33]

## Results

### Study Flow and Participant Characteristics

Of the 651 patients receiving hemodialysis at the six participating facilities, 484 (74.3%) participated in the study. Of these, one patient who was not taking the medication regularly was excluded. Therefore, 483 patients were analyzed.

Table 1 outlines study participant characteristics. The median age of the patients was 71.9 years (IQR, 63.2–78.5), with 48.8% having completed high school and 54.4% reporting an annual household income of <¥3 million. The median dialysis duration was 5.7 years (IQR 2.9LJ10.7 years).

**Table 1.**
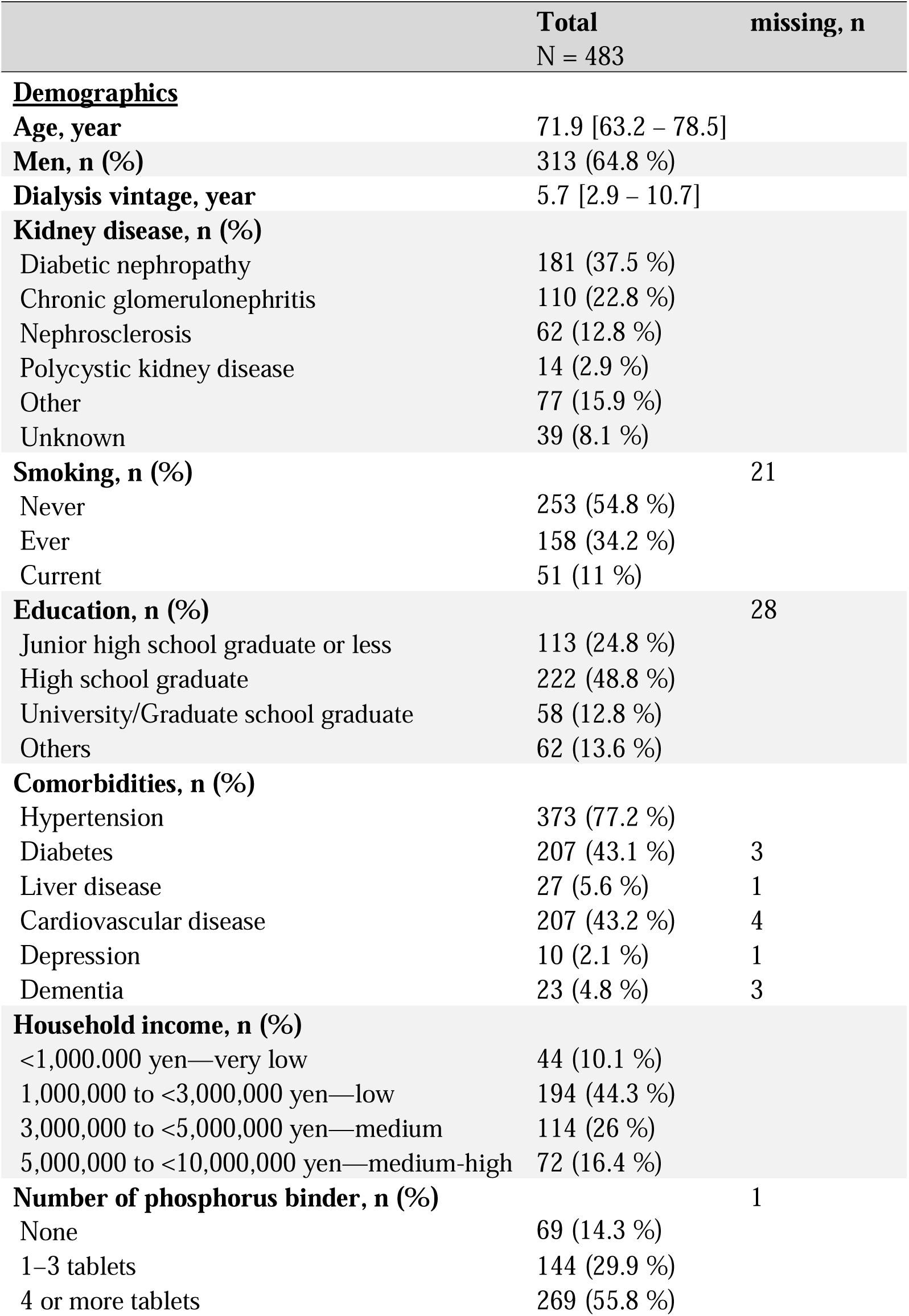
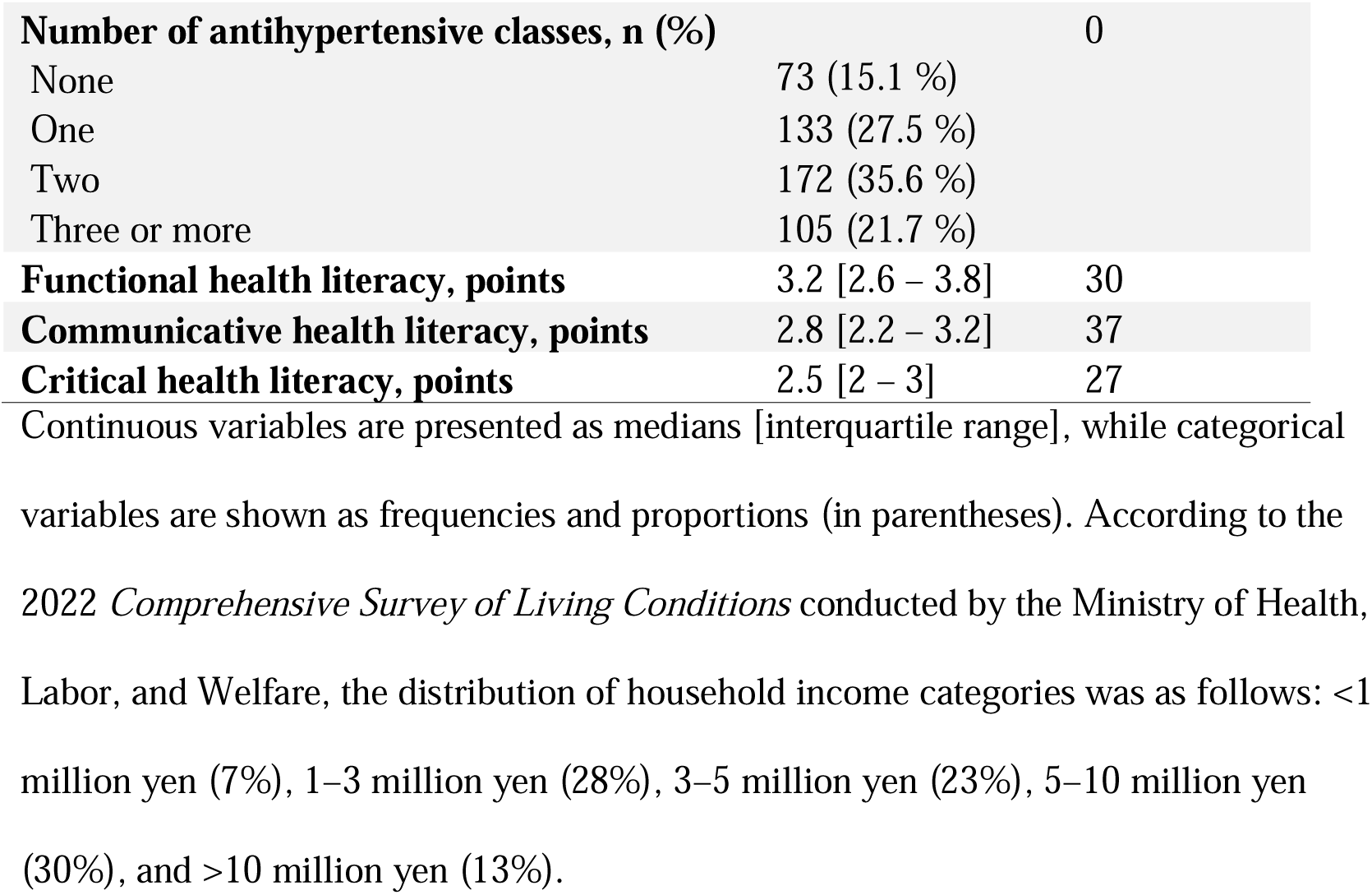
Patient characteristics (N = 483)

### Association between PCC and Trust in Physicians

The median trust in physicians score was 65 (IQR: 55LJ75). Sixty respondents (13.2%) reported having no USC, whereas 396 (66.8%) had USCs, with the median total score on the JPCAT-SF being 60.4 (IQR: 50LJ68.8).

Table 2 outlines the association between the degree of PCC, as measured by the JPCAT-SF total score, and trust in physicians. A dose-dependent association was observed with trust in physicians: with no USC as the reference, the adjusted mean difference increased from 3.57 (95% CI −2.48 to 8.7) for the lowest quartile group to 15.9 (95% CI 9.98 to 21.27) for the highest quartile group.

**Table 2.** Association of trust in physicians with person-centered care.

| Independent variable | Mean difference, point estimate (95% CI) |
| --- | --- |
| **JPCAT-SF total score** |  |
| No Usual Source of Care | Reference |
| Q1 | 3.57 (-2.48 to 8.70) |
| Q2 | **7.40 (1.90 to 12.6)** |
| Q3 | **10.1 (4.91 to 15.1)** |
| Q4 | **15.9 (9.98 to 21.3)** |

Notes: Coefficients $a_1$ to $a_4$ in Figure 1 are described. These coefficients were derived from a general linear model with adjustment for age, sex, dialysis vintage, comorbidities, smoking, number of antihypertensive classes, education, household income, phosphate binder, and multidimensional health literacy. Confidence intervals were calculated using percentile-based bootstrapping with 1,000 resamples. Bold font indicates statistical significance at the 95% confidence level.
The JPCAT-SF total score quartiles: Q1, 0.0–50.0; Q2, 50.0–60.0; Q3, 60.0–70.0; and Q4, 70.0–100.0.
CI, confidence interval; JPCAT-SF, Japanese version of the Primary Care Assessment Tool–Short Form.

Similar dose-response relationships for adjusted mean differences were observed for the JPCAT-SF subdomain scores from the bottom two quartiles to the highest quartile, with some exceptions for the lowest quartile group. (Supplementary Table 1)

### Association of Medication Adherence with PCC and Trust in Physicians

The median ASK-12 total score was 23 (IQR: 19LJ28). Table 3 summarizes the relationships between the ASK-12 total score, the JPCAT-SF total score, and trust in physicians. Total JPCAT-SF scores showed a dose-dependent association with less adherence difficulties: with no USC as the reference, the adjusted mean difference decreased from −3.90 (95% CI −5.95 to −1.75) in the lowest quartile to −4.95 (95% CI −7.21 to −2.62) for the highest quartile.

**Table 3.**
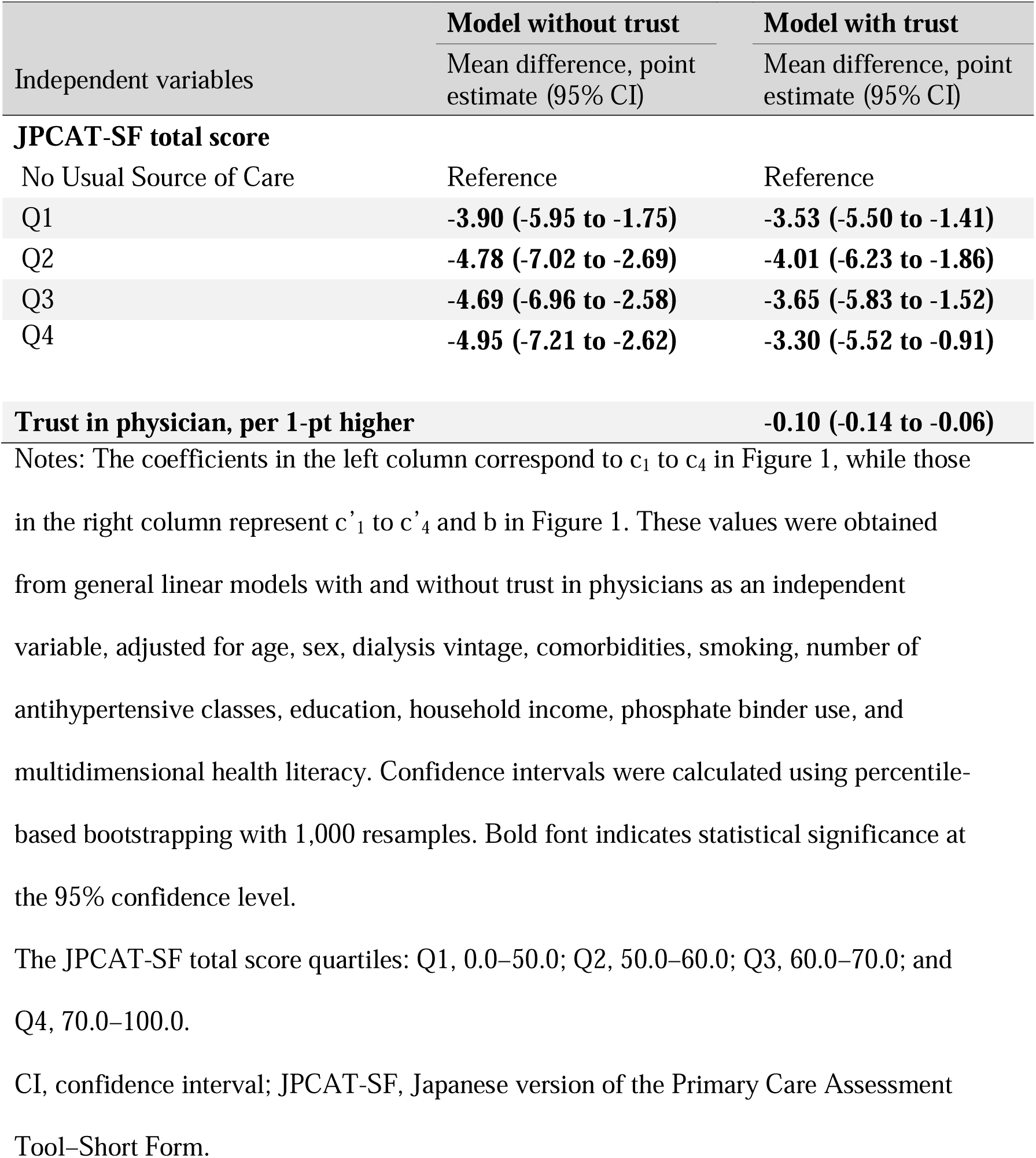
Association of medication adherence score with person-centered care and trust in physicians.

| Independent variables | Model without trust<br>Mean difference, point estimate (95% CI) | Model with trust<br>Mean difference, point estimate (95% CI) |
| --- | --- | --- |
| **JPCAT-SF total score** |  |  |
| No Usual Source of Care | Reference | Reference |
| Q1 | **-3.90 (-5.95 to -1.75)** | **-3.53 (-5.50 to -1.41)** |
| Q2 | **-4.78 (-7.02 to -2.69)** | **-4.01 (-6.23 to -1.86)** |
| Q3 | **-4.69 (-6.96 to -2.58)** | **-3.65 (-5.83 to -1.52)** |
| Q4 | **-4.95 (-7.21 to -2.62)** | **-3.30 (-5.52 to -0.91)** |
| **Trust in physician, per 1-pt higher** |  | **-0.10 (-0.14 to -0.06)** |

Notes: The coefficients in the left column correspond to $c_1$ to $c_4$ in Figure 1, while those
in the right column represent $c'_1$ to $c'_4$ and $b$ in Figure 1. These values were obtained
from general linear models with and without trust in physicians as an independent
variable, adjusted for age, sex, dialysis vintage, comorbidities, smoking, number of
antihypertensive classes, education, household income, phosphate binder use, and
multidimensional health literacy. Confidence intervals were calculated using percentile-
based bootstrapping with 1,000 resamples. Bold font indicates statistical significance at
the 95% confidence level.
The JPCAT-SF total score quartiles: Q1, 0.0–50.0; Q2, 50.0–60.0; Q3, 60.0–70.0; and
Q4, 70.0–100.0.

When adjusted for trust in physicians, the magnitude of the association between the JPCAT-SF total score and medication adherence was smaller, indicating a dose-independent relationship: For example, the adjusted mean difference in the ASK-12 total score was −3.53 (95% CI: −5.5 to −1.41) for the lowest quartile of the JPCAT-SF total score compared to −3.3 (95% CI: −5.52 to −0.91) for the highest quartile. Trust in physicians was also significantly associated with lower adherence difficulties, independent of the JPCAT-SF total score. The adjusted mean difference in the ASK-12 total score was −0.10 (95% CI: −0.14 to −0.06) per 1-point higher trust in physicians.

Similar associations were observed when the JPCAT-SF subdomain scores were treated as exposures (Supplementary Table 2).

### Mediating Effects of Trust in Physicians on the Relationship Between PCC and Medication Adherence

Table 4 and Supplementary Table 3 outline the results of the mediation analyses. The JPCAT-SF total score positively correlated with the total score of medication adherence in a dose-response manner, and trust in physicians partially mediated this relationship, accounting for 16.1% to 33.3% of the correlation from the second quartile to the highest quartile (Table 4).

**Table 4.**
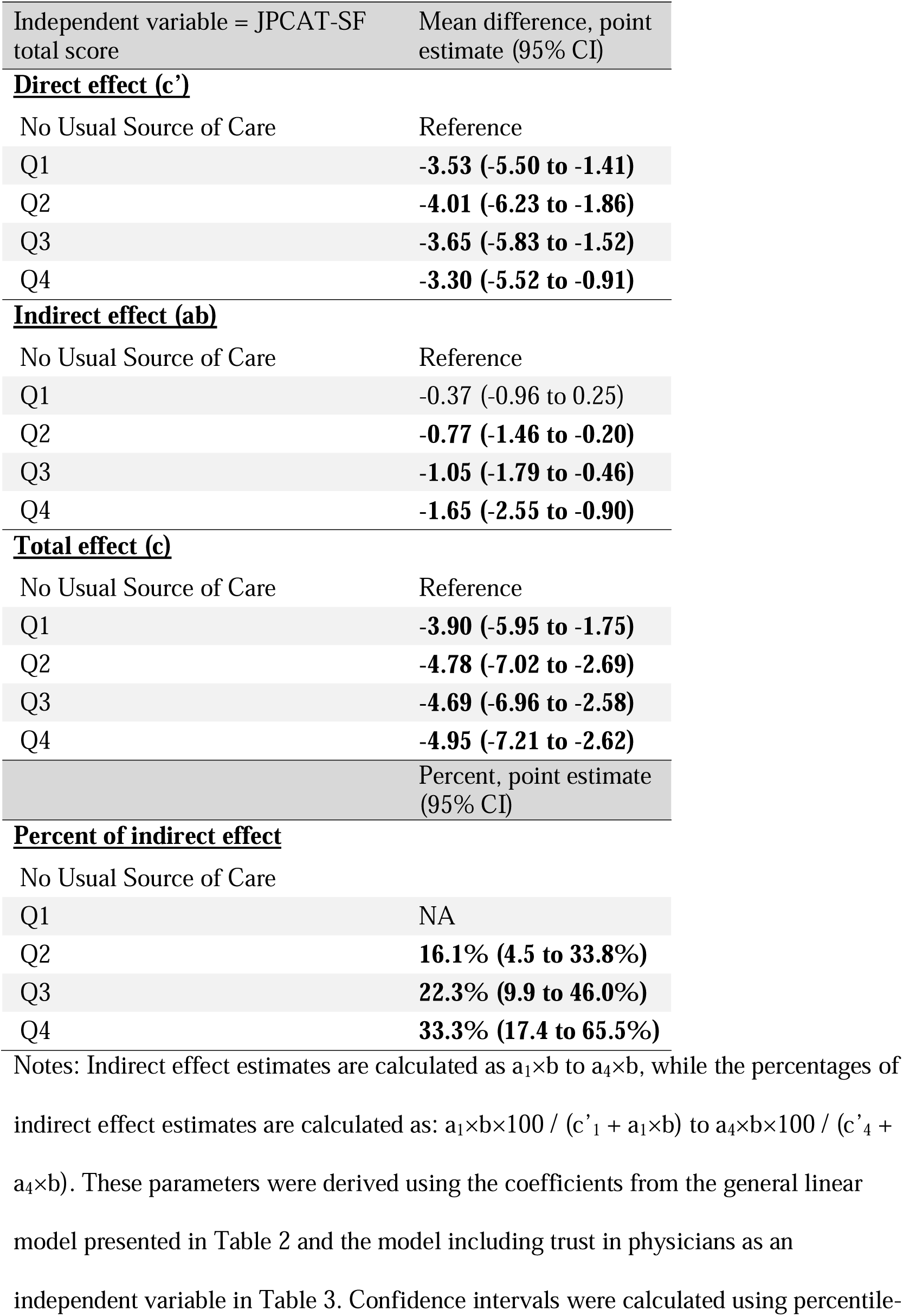
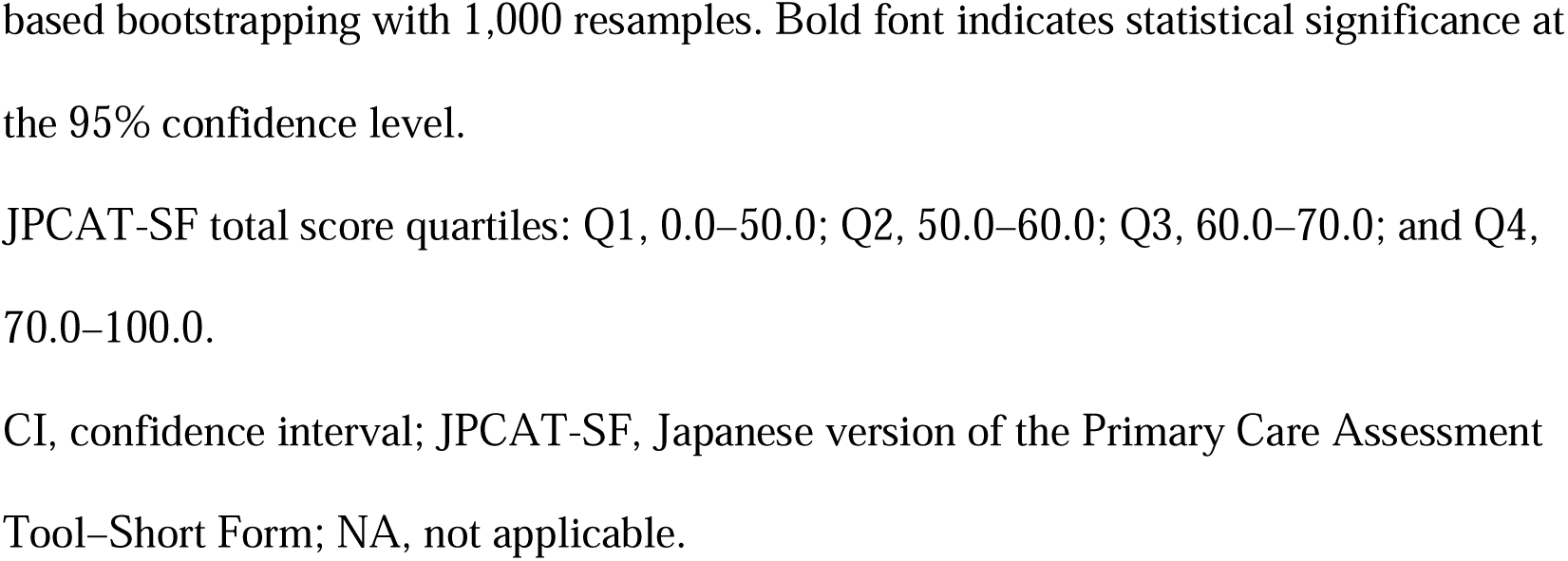
Mediating effect of person-centered care on medication adherence.

Similarly, most of the JPCAT-SF subdomain scores (First Contact, longitudinality, Comprehensiveness [Services Available], Community Orientation) positively correlated with medication adherence total score in a quantity-response manner (Supplementary Table 3). Trust in physicians partially mediated each subdomain correlation in a quantity-response manner, with some exceptions in the lowest and second quartiles.

## Discussion

This study demonstrated that high-quality PCC is positively associated with good medication adherence in a dose-response manner, with trust in physicians mediating this relationship. Notably, the mediating effect of trust in physicians also exhibited a dose-response pattern. These associations remained consistent when individual PCC constructs, such as longitudinality and coordination of care, were analyzed.

The findings of this study reinforce and extend the existing nephrology literature on the importance of PCC, highlighting its role in fostering trust and improving medication adherence. Several systematic reviews of qualitative studies on patients with CKD have suggested PCC approaches to facilitate medication adherence, [8] including continuity in decision-making about medications, respect for patient medication values, [10] and coordination of care. [11] However, despite frequent hypothesis generation, our study is the first to demonstrate that high-quality PCC is associated with improved medication adherence among patients undergoing hemodialysis. Although a quantitative association between high-quality PCC and good medication adherence has been reported among patients with hypertension, the PCC measure used in that study included trust in physicians.[34] As a result, it remains unclear whether the observed adherence was attributable to trust in physicians or independently to the provision of high-quality PCC. The demonstration of a dose-response association between PCC quality and trust in physicians reinforces findings from a previous study that showed a relationship between patient-centered communication and trust in nephrologists among patients with CKD. [18] The emphasis on understanding what matters most to patients with CKD, as reflected by longitudinality, aligns with O’Hare’s argument that such understanding is foundational to PCC, building and strengthening trust in renal medicine. [13] An increase in empathic communication likely fosters a sense of security and trust in physicians, thereby encouraging patients to share their values and concerns regarding medications. [10]

Coordination between nephrologists and other specialists, as reflected in the coordination of care, may help prevent duplicate prescriptions or the prescribing of medications with opposing actions. This coordination also likely reduces discrepancies in medication recommendations among healthcare providers, which can arise from the involvement of multiple physicians rotating shifts at the same dialysis facility—a practice that may be unique to Japan—or from interactions between physicians, nurses, and other co-medical professionals. Moreover, a qualitative study has highlighted that discrepancies in treatment recommendations can impair medication adherence and serve as a key area for improving communication with patients with CKD. [11] Conversely, discontinuity of care has been associated with diminishing trust in the usual physicians of patients, particularly among those with systemic lupus erythematosus. [35]

These findings highlight the importance of researchers and dialysis physicians addressing variability in PCC quality and the role of physician mediation in developing tailored interventions for improving medication adherence. [9,19] In particular, mediation analyses can elucidate the extent to which interventions aimed at enhancing PCC quality improve medication adherence through the mediation of trust in physicians. [36]

The partially mediating role of trust in physicians in the association between PCC and medication adherence highlights the potential to improve adherence through both physician-patient interactions and other channels. [9] Physicians can discuss the benefits and risks of medications in alignment with what matters most to their patients and demonstrate an empathetic understanding of their values regarding medications. [12] Such communication fosters trust and facilitates complex decision-making about medications. [12,37] In addition, patients undergoing hemodialysis spend considerable time with nurses, other healthcare professionals, and physicians. [9] Nurses, in particular, can engage with patients empathetically as whole individuals, supporting their daily lives and addressing adherence issues by organizing medications. Thus, clear role allocation for medication adherence practices at the facility level—often complementary among healthcare providers—may be essential. [38]

Moreover, the finding that PCC quality affects medication adherence independently of physician involvement (a direct effect) underscores the need to improve care quality at the dialysis facility level. For example, ensuring off-hours care, as captured by first contact, could reduce the need for medications related to CKD complications by extending dialysis sessions into off-hours or adding extra sessions when necessary.

Consultations on lifestyle and self-medication, captured by comprehensiveness (service provided), provide patients undergoing dialysis opportunities to address issues such as improving bowel habits. Understanding and addressing these symptoms is a vital role for both nurses and dialysis physicians. For instance, appropriate consultations can improve medication adherence by resolving misconceptions [10] (such as suspecting constipation is caused by medications when it is not), adjusting medications to alternatives that are less likely to cause constipation, or deprescribing unnecessary medications. [38]

This study has several strengths. First, its multicenter design and high participation rate enhance the generalizability of the findings. Second, the use of formal mediation analysis enabled us to quantify the extent to which trust in physicians mediates the association between various aspects of PCC and medication adherence. Third, this study was able to evaluate PCC across multiple subdomains, advancing beyond the qualitative descriptions commonly used in previous studies. [39] This approach provided valuable insights into specific aspects of PCC that may require modification to improve medication adherence.

However, this study has some limitations. First, due to its cross-sectional design, the possibility of reverse causation cannot be excluded. For instance, low medication adherence might result in poorer health status and diminished trust in physicians, potentially undermining the validity of our mediation analyses. Second, medication adherence was assessed through self-reporting, as in previous studies, [9] and did not account for factors such as the number of pills, regimen complexity, or dosage forms. Third, the trust in physicians scale used in this study evaluates trust in physicians generally and may not capture trust in individual physicians. However, in Japan, patients undergoing dialysis are typically attended by multiple physicians rotating in shifts. [9]

In conclusion, our findings suggest that high-quality PCC is associated with improved medication adherence among patients undergoing maintenance hemodialysis. Further, the results indicate that trust in physicians partially mediates the relationship between PCC and medication adherence. Enhancing the quality of individual PCCs and fostering strong physician-patient interactions may be effective strategies to improve medication adherence.

## Authors’ Contributions

Research idea and study design: YK, RI, T. Toida, NK; data acquisition: YK, RI, MU, T. Toishi, AK, MM, TO, YM, TS; data analysis/interpretation: YK, RI, T. Toida, TA, TS, NK; statistical analysis: YK, NK; supervision or mentorship: NK. Each author contributed important intellectual content during manuscript drafting or revision, agreed to be personally accountable for the individual’s contributions, and ensured that questions pertaining to the accuracy or integrity of any portion of the work, even one in which the author was not directly involved, were appropriately investigated and resolved, including documentation in the literature, if appropriate.

## Data Availability

All data produced in the present work are contained in the manuscript.

## Acknowledgments

The authors greatly thank the following researchers, research assistants, and medical staff members for their assistance in collecting the questionnaire-based and clinical information used in this study: Ms. Aki Tairaku (Shin-Yurigaoka General Hospital, Kawasaki-City, Kanagawa); Ms. Takako Saruwatari and Ms. Akiko Kamimura (Kyushu University of Health and Welfare, Nobeoka-City, Miyazaki); Tetuo Ueki, MD, Akio Munakata, MD, Yoshihiko Watanabe, MD (Munakata Clinic, Mobara-City, Chiba); Ms. Yayoi Takanashi, Reiji Masaki, NP, Tomohiko Inoue, MD, Shinnosuke Sugihara, MD, Kanako Nagaoka, MD and Hiroshi Kuji, MD (Kameda Medical Center, Kamogawa-City, Chiba); Kenji Yamaguchi, MD (Awa Regional Medical Center, Tateyama-City, Chiba); and Ms. Miyuki Sato (Fukushima Medical University Hospital, Fukushima-City, Fukushima).

