## Supplementary Files for "Trust in physicians as a mediator of the relationship between person-centered care and medication adherence in patients undergoing hemodialysis: A cross-sectional study"

### Supplementary Item 1. The Adherence Starts with Knowledge 12 scale (ASK-12) ^1,2^

| Instruction sentence | Taking Medicine-What Gets in the Way? Think about all of the medicines you take. Mark one answer for each item below. |
| --- | --- |
| Lifestyles | |
| Question 1 | I forget to take my medicines some of the time. |
| Question 2 | I run out of my medicines because I don’t get refills on time. |
| Question 3 | Taking medicines more than once a day is inconvenient. |
| Attitudes and Beliefs | |
| Question 4 | I feel confident that each one of my medicines will help me. |
| Question 5 | I know if I am reaching my health goals. |
| Help From Others | |
| Question 6 | I have someone whom I can call with questions about my medicines. |
| Talking With Healthcare Team | |
| Question 7 | My doctor/nurse and I work together to make decisions. |
| Taking Medicines | |
| Question 8 | Have you taken a medicine more or less often than prescribed? |
| Question 9 | Have you skipped or stopped taking a medicine because you didn’t think it was working? |
| Question 10 | Have you skipped or stopped taking a medicine because it made you feel bad? |
| Question 11 | Have you skipped, stopped, not refilled, or taken less medicine because of the cost? |
| Question 12 | Have you not had medicine with you when it was time to take it? |
| Response options for questions 1 to 7 | Strongly agree / Agree / Neutral / Disagree / Strongly disagree |
| Response options for questions 8 to 12 | In the last week / In the last month / In the last 3 months / More than 3 months ago / Never |

The English version ^3^ is provided for each item and response.

Questions 1 through 3 constitute the “Inconvenience/Forgetfulness” domain, questions 4 through 7 the “Treatment Beliefs” domain, and questions 8 through 12 the “Behavior” domain.

**Supplementary Item 2. Items and responses for the JPCAT-SF [1]**

Questionnaires in Japanese version are available from the following website (<https://bfffe681-45f7-48c3-aa93-2f67612434a5.filesusr.com/ugd/6c0e9c_7c80f3e3ce6e45eebb5a3aedfb9a2800.pdf>).

| **Instruction sentences** | **(English: “Check the box that best fits each question.”)** |
| --- | --- |
| Question 1 | (English: “When your Primary Care Practice is closed on Saturday and Sunday and you get sick, would someone from there see you the same day?”) |
| Response to Question 1 | (English: “Strongly agree/Somewhat agree/Not sure/Somewhat disagree/Strongly disagree ”) |
| Question 2 | (English: “When your Primary Care Practice is closed and you get sick during the night, would someone from there see you that night?”) |
| Response to Question 2 | (English: “Strongly agree/Somewhat agree/Not sure/Somewhat disagree/Strongly disagree ”) |
| Question 3 | (English: “Does your Primary Care Physician (PCP) know you very well as a person, rather than as someone with a medical problem?”) |
| Response to Question 3 | (English: “Strongly agree/Somewhat agree/Not sure/Somewhat disagree/Strongly disagree ”) |
| Question 4 | (English: “Does your PCP know what problems are most important to you?”) |
| Response to Question 4 | (English: “Strongly agree/Somewhat agree/Not sure/Somewhat disagree/Strongly disagree ”) |
| Question 5 | (English: “Have you ever had a visit to a specialist or special service of any kind?”) |
| Response to Question 5 | (English: “Yes/No or Not sure”) |
| Question 6 | (English: “Did your PCP suggest you go to the specialist or special service?”) |
| Response to Question 6 | (English: “Strongly agree/Somewhat agree/Not sure/Somewhat disagree/Strongly disagree ”) |
| Question 7 | (English: “Did your PCP discuss with you the different places you could have visited to get help with that problem?”) |
| Response to Question 7 | (English: “Strongly agree/Somewhat agree/Not sure/Somewhat disagree/Strongly disagree ”) |
| Question 8 | (English: “Please indicate whether it is available at your PCP’s office. Counselling related to abuse”) |
| Response to Question 8 | (English: “Strongly agree/Somewhat agree/Not sure/Somewhat disagree/Strongly disagree ”) |
| Question 9 | (English: “Please indicate whether it is available at your PCP’s office. Counselling related to personal preferences about end-of-life issues”) |
| Response to Question 9 | (English: “Strongly agree/Somewhat agree/Not sure/Somewhat disagree/Strongly disagree ”) |
| Question 10 | (English: “In visits to your PCP, are any of the following subjects discussed with you? Advice about over-the-counter medications.”) |
| Response to Question 10 | (English: “Strongly agree/Somewhat agree/Not sure/Somewhat disagree/Strongly disagree ”) |
| Question 11 | (English: “In visits to your PCP, are any of the following subjects discussed with you? Advice about medical information in the media: on TV, in the newspaper, etc.”) |
| Response to Question 11 | (English: “Strongly agree/Somewhat agree/Not sure/Somewhat disagree/Strongly disagree ”) |
| Question 12 | (English: “Does your PCP investigate whether the available health care is meeting the needs of the community?”) |
| Response to Question 12 | (English: “Strongly agree/Somewhat agree/Not sure/Somewhat disagree/Strongly disagree ”) |
| Question 13 | (English: “Does your PCP investigate the concerns people have about health problems in your community?”) |
| Response to Question 13 | (English: “Strongly agree/Somewhat agree/Not sure/Somewhat disagree/Strongly disagree ”) |

Each domain consists of the following items:

First contact domain - Questions 1 and 2

Longitudinality domain - Questions 3 and 4

Coordination domain - Questions 5, 6, and 7

Comprehensiveness (services available) domain - Questions 8 and 9

Comprehensiveness (services provided) domain - Questions 10 and 11

Community orientation domain - Questions 12 and 13

**Scoring**

For each item, participants were asked to respond on a 5-point Likert scale ranging from *strongly disagree* to *strongly agree*. Each response was converted to an item score ranging from 0 to 4. The domain scores were calculated by multiplying the average of the item scores in the same domain by 25 (i.e., ranging from 0 to 100), with higher scores indicating better performance. In the coordination domain, which asks about experiences with referrals to a specialist, respondents who had never seen a specialist were given 50 points (the midpoint of all possible scores). The total score was the average of the six domain scores and represented an overall measure of the patient experience of primary care.

### Supplementary Item 3. Description of the concepts of the JPCAT-SF subdomains

The JPCAT-SF is a short version of the original 29-item JPCAT,^1^ which is an adaptation to the Japanese culture of the Primary Care Assessment Tool (PCAT) that was designed to measure the experience of adult patients in primary care.^2^ In an outpatient setting, the JPCAT-SF has been shown to have good internal consistency reliability (Cronbach’s α = 0.77 for the total score, Cronbach’s α > 0.76 for each domain score) and excellent criterion validity (Pearson correlation coefficient with the original 29-item JPCAT and the overall rating for usual care facilities: 0.94 and 0.43, respectively).^3^

First contact

Care is first sought from a primary care provider when a new health or medical need arises. The service should also be accessible and usable by the population as a new need or problem arises.^4^ First contact is closely related to “access to care,” a domain of person-centered care characterized by the timely availability of care that is tailored to the patient.^5^ The JPCAT-SF mainly measures patient experience related to off-hours care in primary care.^4^

Longitudinality

It refers to the longitudinal use of usual sources of care regardless of illness or injury.^4^ Longitudinality is supported by one of the principles for person-centeredness, namely the consideration of the “patient as a unique person,” i.e., the primary care physician’s recognition of the patient’s uniqueness (individual needs, preferences, values, beliefs, concerns, etc.).^5^ The JPCAT-SF mainly measures whether a patient feels that their primary care physician recognizes them as a whole person.^4^

Coordination

The essence of coordination is the availability of information about past and existing problems and services and the recognition of that information in relation to a current care need.^4^ It relates to “coordination and continuity of care,” which is an enabler of person-centered care, i.e., facilitation of care that is well-coordinated and continuous.^5^ The JPCAT-SF mainly measures patient experience regarding past referrals to a specialist.^4^

Comprehensiveness (services available)

It refers to the availability of a wide range of services by a primary care provider and their appropriateness for a spectrum of needs for all but the most uncommon problems.^4^

Under “services available,” the JPCAT-SF mainly measures whether a patient feels they can receive care for mental health, dementia, and advanced care planning, if necessary.^4^

Comprehensiveness (services provided)

It includes appropriate advice on daily lifestyle habits, self-medication, and health literacy.^4,6^ It is underpinned by patient empowerment, an activity of person-centeredness, in which a primary care physician recognizes and actively supports a patient's ability and responsibility to self-manage their illness.^5^ The JPCAT-SF mainly measures patient experience in terms of whether they received such appropriate advice.

Community orientation

It refers to care that is delivered in the context of the community^4^ and is considered as a derivative domain of principles of primary care.^1^

The JPCAT-SF mainly measures patient experience regarding home visits and whether a patient feels that their primary care physician is interested not only in their individual health problem but also in problems in the community.^4^

### Supplementary Item 4: A brief description of the definition of covariates

Cardiovascular disease was defined as a history of myocardial infarction, angina pectoris, heart failure, cerebrovascular disease, or peripheral vascular disease. Hypertension was defined as taking antihypertensive medications regularly or having a systolic blood pressure ≥ 140 mmHg or at the first dialysis session each week (typically before starting dialysis on Monday or Saturday). A prescription of antihypertensives was considered present if any of the following was prescribed, where each antihypertensive agent was categorized into five classes for analysis: (i) calcium channel blockers, (ii) angiotensin-converting enzyme inhibitors, angiotensin II receptor blockers, angiotensin receptor neprilysin inhibitors, (iii) alpha-beta blockers, beta-blockers, (iv) alpha-blockers, and (v) central sympatholytic agents. The pill count for the phosphate binders was calculated if any of the following were prescribed: calcium-based, sevelamer, bixalomer, lanthanum, sucroferric oxyhydroxide, or ferrous citrate hydrate.

### Supplementary Figure 1. Conceptual framework for this study.

Analysis 1: Association between trust in physicians and person-centered care.

Analysis 2: Association between person-centered care and medication adherence, modeled both with and without trust in physicians.

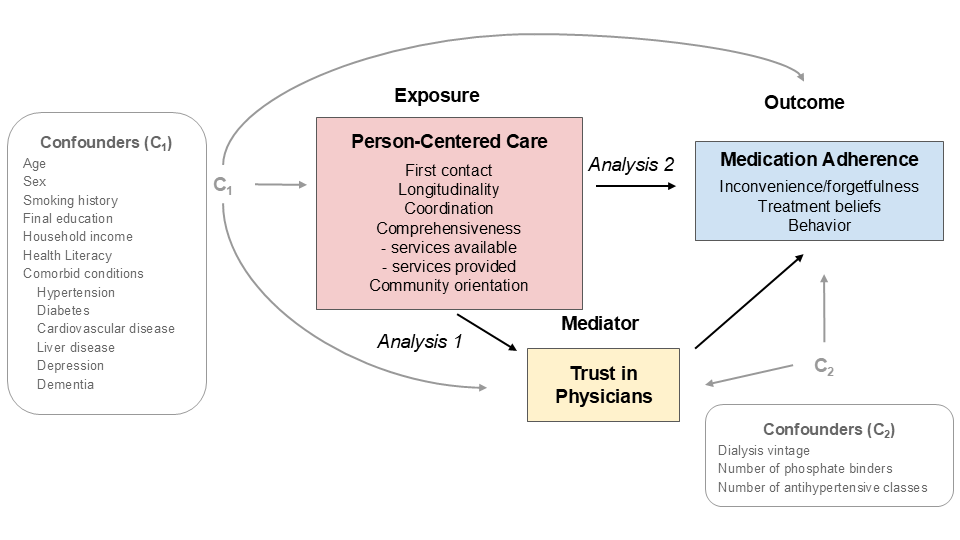

### Supplementary Table 1. Association of trust in physicians with person-centered care

|  | **Mean difference, point estimate (95% CI)** | | | | | | | | | | |
| --- | --- | --- | --- | --- | --- | --- | --- | --- | --- | --- | --- |
| Independent variable | First Contact |  | Longitudinality |  | Coordination |  | Comprehensiveness (Services Available) |  | Comprehensiveness (Services Provided) |  | Community Orientation |
| **JPCAT-SF subdomain score** | |  |  |  |  |  |  |  |  |  |  |
| No Usual Source of Care | Reference |  | Reference |  | Reference |  | Reference |  | Reference |  | Reference |
| Q1 | **6.79 (1.32 to 12.2)** |  | -2.35 (-8.88 to 4.03) |  | **7.87 (2.50 to 12.7)** |  | 4.46 (-1.46 to 10.1) |  | **11.9 (5.93 to 16.3)** |  | 1.64 (-3.72 to 6.80) |
| Q2 | **5.70 (0.56 to 10.9)** |  | **4.41 (-0.89 to 9.26)** |  | **5.90 (0.28 to 11.0)** |  | **6.70 (1.40 to 11.4)** |  | **5.10 (-0.15 to 9.70)** |  | **7.30 (1.93 to 12.1)** |
| Q3 | **10.5 (5.27 to 15.48)** |  | **10.9 (5.69 to 15.7)** |  | **9.77 (4.00 to 14.9)** |  | **9.72 (4.80 to 15.0)** |  | **9.07 (2.84 to 14.0)** |  | **11.9 (7.10 to 16.8)** |
| Q4 | **12.7 (6.35 to 17.65)** |  | **16.6 (11.1 to 21.8)** |  | **12.0 (6.17 to 17.2)** |  | **12.8 (6.62 to 18.5)** |  | **10.1 (3.46 to 15.1)** |  | **18.7 (12.3 to 24.4)** |

Coefficients were obtained from general linear models adjusted for age, sex, dialysis vintage, comorbidities, smoking, number of antihypertensive classes, education, household income, phosphate binder use, and multidimensional health literacy. Confidence intervals were calculated using percentile-based bootstrapping with 1,000 resamples. Significant associations at the 95% confidence level are shown in bold.

JPCAT-SF Subdomain Score Quartiles:

- First Contact: Q1, 0.0–37.5; Q2, 50.0–62.5; Q3, 75.0; Q4, 87.5–100.0
- Longitudinality: Q1, 0.0–37.5; Q2, 50.0–62.5; Q3, 75.0; Q4, 87.5–100.0
- Coordination: Q1, 0.0–50.0; Q2, 62.5–75.0; Q3, 87.5; Q4, 100.0
- Comprehensiveness (Services Available): Q1, 0.0–37.5; Q2, 50.0; Q3, 62.5–75.0; Q4, 87.5–100.0
- Comprehensiveness (Services Provided): Q1, 0.0; Q2, 12.5–25.0; Q3, 37.5–50.0; Q4, 62.5–100.0
- Community Orientation: Q1, 0.0–37.5; Q2, 50.0; Q3, 62.5–75.0; Q4, 87.5–100.0

*JPCAT-SF, Japanese version of the Primary Care Assessment Tool–Short Form*

### Supplementary Table 2. Association of medication adherence score with person-centered care and trust in physicians

| **Mean difference, point estimate (95% CI)** | | | | | | | | | | | |
| --- | --- | --- | --- | --- | --- | --- | --- | --- | --- | --- | --- |
| Independent variables | First Contact |  | Longitudinality |  | Coordination |  | Comprehensiveness (Services Available) |  | Comprehensiveness (Services Provided) |  | Community Orientation |
| **Model without trust** | |  |  |  |  |  |  |  |  |  |  |
| No Usual Source of Care | Reference |  | Reference |  | Reference |  | Reference |  | Reference |  | Reference |
| Q1 | **-4.19 (-6.58 to -2.10)** |  | -2.44 (-4.97 to 0.23) |  | **-4.72 (-6.81 to -2.78)** |  | **-3.76 (-6.37 to -1.44)** |  | **-5.76 (-7.79 to -3.77)** |  | **-3.22 (-5.66 to -1.15)** |
| Q2 | **-4.46 (-6.70 to -2.48)** |  | **-4.44 (-6.56 to -2.42)** |  | **-3.80 (-5.97 to -1.69)** |  | **-4.12 (-6.23 to -1.99)** |  | **-3.93 (-6.05 to -1.85)** |  | **-4.69 (-6.79 to -2.66)** |
| Q3 | **-4.55 (-6.73 to -2.35)** |  | **-4.46 (-6.57 to -2.23)** |  | **-4.55 (-6.80 to -2.67)** |  | **-4.80 (-6.90 to -2.69)** |  | **-4.22 (-6.25 to -2.06)** |  | **-4.63 (-6.96 to -2.59)** |
| Q4 | **-4.83 (-7.20 to -2.48)** |  | **-5.67 (-7.77 to -3.46)** |  | **-4.73 (-6.98 to -2.67)** |  | **-5.35 (-7.73 to -3.22)** |  | **-4.21 (-6.38 to -1.75)** |  | **-6.29 (-8.78 to -3.95)** |
| **Model with trust** | |  |  |  |  |  |  |  |  |  |  |
| No Usual Source of Care | Reference |  | Reference |  | Reference |  | Reference |  | Reference |  | Reference |
| Q1 | **-3.47 (-5.74 to -1.40)** |  | **-2.67 (-4.98 to -0.18)** |  | **-3.90 (-5.98 to -1.97)** |  | **-3.29 (-5.88 to -1.09)** |  | **-4.56 (-6.65 to -2.62)** |  | **-3.06 (-5.40 to -0.98)** |
| Q2 | **-3.85 (-5.99 to -1.96)** |  | **-4.00 (-6.01 to -2.07)** |  | **-3.19 (-5.39 to -1.13)** |  | **-3.43 (-5.59 to -1.40)** |  | **-3.41 (-5.55 to -1.40)** |  | **-3.96 (-5.97 to -1.97)** |
| Q3 | **-3.43 (-5.65 to -1.34)** |  | **-3.37 (-5.46 to -1.13)** |  | **-3.54 (-5.74 to -1.67)** |  | **-3.79 (-5.86 to -1.80)** |  | **-3.30 (-5.37 to -1.36)** |  | **-3.44 (-5.73 to -1.47)** |
| Q4 | **-3.48 (-5.87 to -1.20)** |  | **-4.01 (-6.16 to -1.82)** |  | **-3.48 (-5.81 to -1.39)** |  | **-4.02 (-6.36 to -1.95)** |  | **-3.19 (-5.39 to -0.85)** |  | **-4.42 (-6.94 to -2.10)** |
| Trust in physician, per 1-pt higher | **-0.11 (-0.14 to -0.07)** |  | **-0.10 (-0.13 to -0.06)** |  | **-0.10 (-0.14 to -0.06)** |  | **-0.10 (-0.13 to -0.06)** |  | **-0.10 (-0.13 to -0.06)** |  | **-0.10 (-0.14 to -0.06)** |

Coefficients were derived from general linear models with and without trust in physicians as an independent variable. Adjustments were made for age, sex, dialysis vintage, comorbidities, smoking status, number of antihypertensive classes, education, household income, phosphate binder use, and multidimensional health literacy. Confidence intervals were calculated using percentile-based bootstrapping with 1,000 resamples. Significant associations at the 95% confidence level are shown in bold.

**JPCAT-SF Subdomain Score Quartiles:**
(Same as in Supplementary Table 1)

### Supplementary Table 3. Mediating effect of person-centered care on medication adherence

|  | **Mean difference, point estimate (95% CI)** | | | | | |
| --- | --- | --- | --- | --- | --- | --- |
| Independent variables = JPCAT-SF subdomain scores | First Contact | Longitudinality | Coordination | Comprehensiveness (Services Available) | Comprehensiveness (Services Provided) | Community Orientation |
| **Direct effect** |  |  |  |  |  |  |
| No Usual Source of Care | Reference | Reference | Reference | Reference | Reference | Reference |
| Q1 | **-3.47 (-5.74 to -1.40)** | **-2.67 (-4.98 to -0.18)** | **-3.90 (-5.98 to -1.97)** | **-3.29 (-5.88 to -1.09)** | **-4.56 (-6.65 to -2.62)** | **-3.06 (-5.40 to -0.98)** |
| Q2 | **-3.85 (-5.99 to -1.96)** | **-4.00 (-6.01 to -2.07)** | **-3.19 (-5.39 to -1.13)** | **-3.43 (-5.59 to -1.40)** | **-3.41 (-5.55 to -1.40)** | **-3.96 (-5.97 to -1.97)** |
| Q3 | **-3.43 (-5.65 to -1.34)** | **-3.37 (-5.46 to -1.13)** | **-3.54 (-5.74 to -1.67)** | **-3.79 (-5.86 to -1.80)** | **-3.30 (-5.37 to -1.36)** | **-3.44 (-5.73 to -1.47)** |
| Q4 | **-3.48 (-5.87 to -1.20)** | **-4.01 (-6.16 to -1.82)** | **-3.48 (-5.81 to -1.39)** | **-4.02 (-6.36 to -1.95)** | **-3.19 (-5.39 to -0.85)** | **-4.42 (-6.94 to -2.10)** |
| **Indirect effect** |  |  |  |  |  |  |
| No Usual Source of Care | Reference | Reference | Reference | Reference | Reference | Reference |
| Q1 | **-0.72 (-1.36 to -0.13)** | 0.23 (-0.42 to 0.92) | **-0.81 (-1.37 to -0.25)** | -0.46 (-1.09 to 0.14) | **-1.20 (-1.80 to -0.56)** | -0.16 (-0.70 to 0.39) |
| Q2 | **-0.61 (-1.25 to -0.06)** | -0.44 (-1.00 to 0.09) | **-0.61 (-1.15 to -0.03)** | **-0.70 (-1.26 to -0.15)** | -0.52 (-1.04 to 0.02) | **-0.73 (-1.31 to -0.19)** |
| Q3 | **-1.12 (-1.81 to -0.52)** | **-1.09 (-1.72 to -0.45)** | **-1.01 (-1.65 to -0.41)** | **-1.01 (-1.64 to -0.45)** | **-0.92 (-1.52 to -0.28)** | **-1.19 (-1.88 to -0.59)** |
| Q4 | **-1.35 (-2.14 to -0.62)** | **-1.65 (-2.41 to -0.85)** | **-1.24 (-1.91 to -0.54)** | **-1.33 (-2.04 to -0.57)** | **-1.02 (-1.67 to -0.33)** | **-1.86 (-2.80 to -0.97)** |
| **Total effect** |  |  |  |  |  |  |
| No Usual Source of Care | Reference | Reference | Reference | Reference | Reference | Reference |
| Q1 | **-4.19 (-6.58 to -2.10)** | -2.44 (-4.97 to 0.23) | **-4.72 (-6.81 to -2.78)** | **-3.76 (-6.37 to -1.44)** | **-5.76 (-7.79 to -3.77)** | **-3.22 (-5.66 to -1.15)** |
| Q2 | **-4.46 (-6.70 to -2.48)** | **-4.44 (-6.56 to -2.42)** | **-3.80 (-5.97 to -1.69)** | **-4.12 (-6.23 to -1.99)** | **-3.93 (-6.05 to -1.85)** | **-4.69 (-6.79 to -2.66)** |
| Q3 | **-4.55 (-6.73 to -2.35)** | **-4.46 (-6.57 to -2.23)** | **-4.55 (-6.80 to -2.67)** | **-4.80 (-6.90 to -2.69)** | **-4.22 (-6.25 to -2.06)** | **-4.63 (-6.96 to -2.59)** |
| Q4 | **-4.83 (-7.20 to -2.48)** | **-5.67 (-7.77 to -3.46)** | **-4.73 (-6.98 to -2.67)** | **-5.35 (-7.73 to -3.22)** | **-4.21 (-6.38 to -1.75)** | **-6.29 (-8.78 to -3.95)** |
|  | **Percent, point estimate (95% CI)** | | | | | |
| **Percent of indirect effect** |  |  |  |  |  |  |
| No Usual Source of Care |  |  |  |  |  |  |
| Q1 | **17.3% (3.8 to 39.6%)** | NA | **17.3% (5.4 to 32.4%)** | NA | **20.9% (9.6 to 35.3%)** | NA |
| Q2 | **13.7% (1.4 to 30.2%)** | NA | **16.1% (0.9 to 37.8%)** | **16.9% (3.4 to 38.0%)** | NA | **15.6% (3.9 to 31.1%)** |
| Q3 | **24.6% (11.1 to 49.3%)** | **24.4% (10.5 to 51.7%)** | **22.2% (9.1 to 41.0%)** | **21.1% (9.4 to 40.7%)** | **21.7% (6.6 to 44.7%)** | **25.7% (12.3 to 48.7%)** |
| Q4 | **28% (12.8 to 53.4%)** | **29.2% (14.5 to 50.5%)** | **26.3% (11.5 to 50.7%)** | **24.9% (10.9 to 44.7%)** | **24.2% (8.2 to 57.1%)** | **29.6% (14.6 to 51.9%)** |

Mediation analysis was performed using coefficients from the general linear models presented in Supplementary Table 1 and the models incorporating trust in physicians in Supplementary Table 2. Confidence intervals were calculated using percentile-based bootstrapping with 1,000 resamples. Significant mediation effects at the 95% confidence level are shown in bold.

**JPCAT-SF Subdomain Score Quartiles:**
(Same as in Supplementary Table 1)

*JPCAT-SF, Japanese version of the Primary Care Assessment Tool–Short Form; NA, not applicable*
